# An Exploration of the Costs of Family and Group Conferencing Pathways in Adult Social Care and Mental Health: A Mixed Methods Approach

**DOI:** 10.1101/2025.06.07.25329196

**Authors:** Lefan Liu, Jerry Tew, Sharanya Mahesh, Philip Kinghorn

## Abstract

**Context:** Family and Group Conferencing (FGC) is a strengths-based approach to social work, originating from New Zealand and now used internationally. Previous research on FGC has focused largely on the context of children’s services but, FGC also aligns with the principle of the Care Act in England to prevent, reduce or delay the need for long-term (and potentially costly) adult care services. Limited previous research has tended to explore potential cost savings associated with FGC, without accounting for the cost of the intervention itself, risking biased results.

**Objective:** This paper aims to identify resource use and associated monetary costs associated with FGC services in English adult social care and mental health settings.

**Methods:** Framework development was informed by previously published work establishing programme theory for FGC, extended by expert opinion and published sources of monetary costs. The framework used scenario-based analysis and a bottom-up costing approach, with sensitivity analysis.

**Results:** Estimated costs of conducting a standard full FGC (excluding referral) range from £1,455 to £2,043 (adjusted from 2022-2023 to 2025 prices) from a local authority and National Health Service (NHS) perspective. Costs can vary depending on the involvement of an advocate or interpreter, network size and the complexity of issues being addressed.

**Discussion:** Higher staff costs in the UK account for slightly higher intervention costs in an NHS mental health than in an adult social care context.

**Conclusion:** Reallocating scarce public resources with the intention of preventing, reducing or delaying use of costly future care must be evidence-based as pressures build to meet acute needs. Accurate per-case costing of FGC is a necessary preliminary step towards exploring the cost-effectiveness of FGC. A full economic evaluation will account for costs, outcomes, and alternative options (uses of limited resources).

**Ethics Statement:** Using the UK Health Research Authority (HRA) screening tool, it was determined that the wider research project did not constitute research for which HRA approval was necessary, and therefore ethical approval (for the wider research project) was obtained from the University of Birmingham Humanities and Social Sciences Ethical Review Committee (ERN_22-0818). Research reported in this paper did not involve any human participants, or the processing or analysis of any human/personal data. Our research was informed by published sources (including grey literature) and expert opinion from practitioners partnering with us to deliver the research.

## 1 Introduction

Originating in New Zealand in the 1980s, Family and Group Conferencing (FGC) is a family-led decision-making model that is widely used internationally within children and family social services [1]. More recently, the approach has been offered within adult and mental health service contexts and fits within the broader family of strengths-based and preventative approaches [2].

The fundamental philosophy of a strengths-based approach is that strengths exist within all individuals, families and communities, and they are the ‘experts’ on their own challenges and issues [3, 4]. FGC offers a voluntary and inclusive approach in which people can plan for their care and support on their terms – and which can mobilise the strengths and resources that may potentially exist within their family and social networks. It offers an opportunity for family, friends and neighbours to come together with a person to devise their own plan for how best to arrange their care, support and safety. Plans made in this way can be more acceptable both to the person and those involved in their support. They can also reduce the likelihood that responsibility for care and support will fall solely onto one network member. It is widely acknowledged that this approach can reduce the reliance on external resources and helps in the early identification and prevention of potential problems [5, 6].

The Care Act (2014), which applies to local authorities (councils) in England, emphasizes the importance of prevention to minimize the need for formal, long-term care and support for both adults and their carers. The goal is to prevent, reduce, or delay care needs while maximizing individuals’ independence and wellbeing for as long as possible [7]. Since the introduction of the Care Act, there has been growing awareness and interest in Adult Family and Group Conferencing (FGC), as the FGC “way of working” aligns closely to the principles of the Care Act [4, 8].

Use of FGC is not restricted to the contexts of adult social care and children’s services, and is also used within health services. The UK Secretary of State for Health and Social Care recently announced an aspiration to make “Three Shifts” within the National Health Service (NHS), as part of a 10-year health plan, designed to meet the changing needs of a changing population. Two such shifts, which FGC could potentially help facilitate, are: (i) from merely treating sickness to preventing it; and (ii) moving care from hospitals to communities.

Potential cost saving through delaying or preventing intense forms of care services is likely to be a key motivator for public sector organisations funding FGC services, especially in the context of current budget pressures facing local authorities and the NHS in England and challenges meeting demand for long-term care internationally. Quantifying such potential cost savings could help to inform decisions about the allocation of scarce resources. However, there may also be an expectation that FGC will improve outcomes for individuals and families and economic evaluation offers a formal framework within which to compare both the costs and outcomes associated with alternative approaches (such as FGC and standard practice without FGC).

A cost-benefit analysis of an FGC service versus business as usual (BAU), in the context of children’s services and domestic abuse, was conducted in Leeds [9]. The analysis indicated that providing an FGC service is marginally more expensive than existing services, but on average, BAU families spent longer in the social care system, meaning that FGC was associated with savings estimated at £755 per family.

There is no such full economic analysis of FGC in the context of adult social care or NHS mental health services. Research has been limited to quantifying the cost saving associated with reduced residential care, domiciliary care, and care management time, and this has been estimated at between £1,579 per FGC over 12 months [8] in the context of adult safeguarding and £7,000 per FGC over a two-year period [10] in the broader context of adult social care. However, these studies focus on cost savings from the perspective of statutory services and do not account for the cost of delivering FGC as an intervention, changes in the provision of unpaid care nor changes in well-being. There cannot be confident claims of cost saving if the costs of providing FGC services are not accounted for, particularly as cost savings identified in the literature so far can be of relatively modest magnitude.

This is therefore an important gap in the current literature, which we address by presenting detailed, systematic and transparent bottom-up costing of plausible and common FGC pathways. By pathways, we mean the structured processes and procedures followed in implementing FGC models within adult social care and mental health settings. Our use of expert opinion, and the foundation of our pathways in previously identified programme theory, represent novel aspects of our work and are effective ways of circumventing the challenge and complexity of data collection in social care contexts specifically. We account for variations in practice through sensitivity analysis, and account for the fact that not all referrals will lead to a full FGC conference [8, 11].

## 2 Methods

Work reported in this paper forms part of a wider programme of research exploring how Family and Group Conferencing works and what difference it can make to people’s lives and draws on earlier work using survey, interviews and a deliberative forum to gain a contextual understanding of the operation of FGC services [11, 12]. The involvement of people with lived experience was central to the development of the programme theory and was facilitated through both peer researchers with lived experience (who supported literature reviews, interviews and subsequent analysis) and an independent Lived Experience Advisory Panel (LEAP) [11, 12]. The LEAP has guided every aspect of the research, over the full duration of the wider research programme (i.e. beyond development of the programme theory). Results and progress have also been discussed at regular Research and Practice Network meetings.

Our methods here follow three steps: mapping out pathways which detail each stage of the FGC process (Step 1); the identification and quantification of resources used at each stage of the FGC pathway (step 2); and attributing monetary costs to resources used (step 3). Sensitivity analysis explores the impact of changing key assumptions and parameters.

There was no involvement by human research participants and no processing or analysis of personal data. For the wider project (of which, work reported here is one component), ethical approval was obtained from the University of Birmingham Humanities and Social Sciences Ethical Review Committee (ERN_22-0818).

### 2.1. Step 1: Pathway Development

Plausible FGC pathways were directly informed by the existing literature [11, 12]. All pathways cover three characterising stages of FGC: Preparation, Conference, and Review. Furthermore, all pathways were developed from three perspectives: the central person (or service user) around whose needs the FGC is to be convened, network members and the public sector (local authority (LA) and NHS). A base case pathway (Scenario A) reflects all core elements of an FGC process (elements which would need to be included in order to categorise the intervention as being an FGC and align with best practice). Elements which might be added in some cases, dependent on needs and specific circumstances (such as the use of an advocate or an interpreter), are added in additional scenarios to generate a range of values, reflecting plausible variation in practice.

### 2.2 Step 2: Identifying and Quantifying Resource Use

Parameters relating to resource use (step 2) were informed both by previously published research and by expert opinion obtained from FGC leads/managers and practitioners, working across two local authority FGC services and one NHS mental health FGC service; these were established FGC services with geographical spread across England. Informed by this expert opinion and previous research findings, we made several base case assumptions to decide which resources will plausibly be used, and in which quantities, in a typical adult social care setting. The same assumptions apply to mental health settings unless stated otherwise.

We allocate LA/NHS resource use to one of two categories, staff and non-staff, and summarise the quantity of resource use ‘consumed’ at each stage of the FGC process. Staff resources are measured in hours worked. Non-staff resources include travel, food, and venue hire. We also list time commitment for service user and network members at each stage of the FGC so we can estimate the opportunity cost of their time in Step 3.

### 2.3 Step 3: Attributing Monetary Values

In step 3, we conducted bottom-up costing for each of the different FGC pathways (using Scenario A consistently as the base case). To calculate the monetary costs, we assign a value (price) to the resources identified and quantified in step 2, based on several assumptions developed through discussions with the wider project team and FGC practitioners (principally regarding the seniority and salary band of professional staff). We categorized monetary costs from the LA/NHS perspective into staff costs and non-staff costs, according to the type of resource utilized.

For staff costs, we used unit costs (prices) from the Personal Social Services Research Unit (PSSRU) 2022/23 [13]. Mapping to costs from PSSRU has the advantage that non-salary costs associated with employment are accounted for (such as pension and national insurance contributions). Whenever possible, unit costs from PSSRU are used which include qualifications; this approach ensures that training and education costs are accounted for thus reflects the full societal cost of skilled labour. To maintain consistency, we use the 2022/23 price levels for other resources wherever possible. To categorize the monetary costs from the perspectives of central persons and network members, we valued their time based on a shadow wage rate, a common approach used to estimate the opportunity cost of persons’ time when engaging in health or social care services [14]. We assume the opportunity cost for both the person and network members is equivalent to the 2023 UK National Living Wage. This rate is currently the same as the National Minimum Wage for those aged 23 and over [15].

The estimated staff and non-staff costs were then cumulatively summed for each stage of the full FGC service, providing a comprehensive breakdown of the financial implications of an FGC service, primarily from the LA/NHS perspective, but also reporting costs incurred by the person and their network. In sensitivity analysis, we explore the impact of assumptions (such as network size and conference duration) on the overall cost estimates.

Final cost amounts for each pathway (reflecting 2022/2023 prices) are adjusted to March 2025 prices using the Bank of England Inflation Calculator [16].

## 3. Results

### 3.1 FGC Pathways

Figure 1 outlines the three common stages of a basic FGC (the broad pathway). Below Figure 1, we begin to characterise and specify greater detail for each stage.

**Figure 1.**
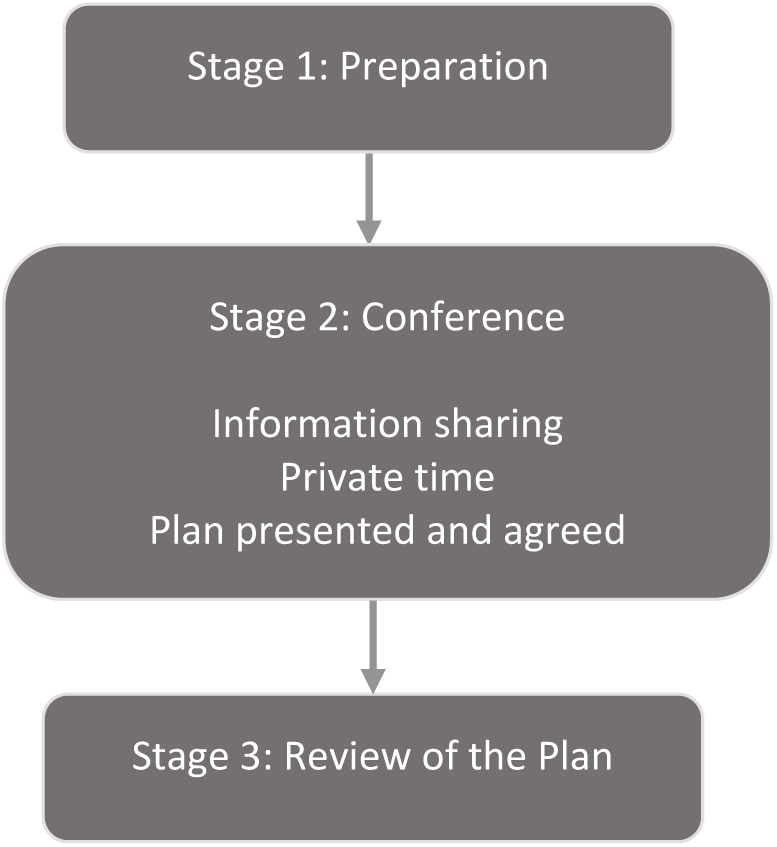
An outline of the basic FGC process

#### Preparation (Stage 1)

The FGC coordinator engages with the central person and invites each network member to participate, establishing what they each hope to achieve from the process. At this stage, the coordinator would help the person to create an invitation list for the FGC, including supporters like family, friends, and any service providers they wish to involve, along with the referrer. Parallel conversations are held with the referrer and other professionals involved to ascertain any issues, such as safeguarding, or other non-negotiables.

#### Conference (Stage 2)

The conference usually takes place at a community venue or the person’s home. At the beginning of the Conference, the coordinator sets (reads out) the pre-decided agenda as well as the ground rules for the Conference. The Conference begins with information sharing, where the coordinator facilitates a conversation between the person, network members and invited professionals. Following this, the person and network members have ‘private time’ to devise a plan independently. Finally, they share the plan (either directly or via the coordinator) and consult with relevant professionals and agencies to ensure the necessary services can be provided and that any professional concerns, such as safeguarding, are addressed.

#### Review (Stage 3)

The coordinator helps assess how the plan is working and adapt it as necessary. A minimum of one review is usually offered, with the option for additional reviews or a further FGC to be offered as appropriate, particularly if circumstances change. Once the review is completed, the FGC referral is closed.

A necessary preliminary stage will always be referral, where a referrer identifies potential suitability for FGC, and the FGC manager or team lead reviews it to ensure it meets the referral criteria. This referral stage is included in some previous evaluations [1]. In our analysis, the referral stage is excluded from base case FGC cost calculations to facilitate a like-for-like comparison across different contexts, because there will be significant variation in the role title and seniority of professionals making referrals. However, we estimate referral costs separately and present separate costs associated specifically with referral.

Among the FGC cases observed by our advising FGC practitioners, approximately 14% of referrals who are accepted into FGC services do not progress to the (full) Conference stage. This is therefore a source of variation in the FGC process and is represented in a separate scenario in the following section (Section 3.2).

### 3.2 Identifying and measuring resource use

Five plausible scenarios emerge from this step, with Scenario A reflecting, wherever possible, the most typical FGC case, and alternative scenarios reflecting variation from that typical base case. Assumptions (A1-A9), underpinning all five scenarios, are as follows:

#### A1 Duration of stages

We assume the FGC coordinator spends significant time on preparation, estimated at 10 hours. FGC practitioners reported that this preparation depends on the size of network and can take up to 20-hours in some cases, although our assumption of 10 hours reflects a typical network size (see A8). The Conference itself is assumed to last for four hours, and the review meeting for two hours. For simplicity, we assume the review meeting will take place online.

#### A2 Involvement of professional staff at the Preparation and Conference Stages

In addition to the FGC Coordinator, other service providers (for example, a representative of the service which made the referral or housing services) may be invited to join the conference by the network (or coordinator) [1]. We assume that two professional staff members attend (in addition to the FGC coordinator), based on the average number of professional staff reported to attend an FGC Conference by our advising FGC practitioners. In addition, we assume that each additional professional spends time (30 minutes) communicating with the coordinator at the preparation stage.

#### A3 Involvement of an advocate at the Conference

For our base case analysis (Scenario A), we assume no professional advocate attends the conference, but we include an advocate in our analysis of an alternative scenario (Scenario B). Advocacy is typically provided when the central person (or a network member) may have a learning disability or cognitive impairment, or may otherwise need support to express their views as part of the Conference process [1]. Advocacy may be provided by an advocacy service or, in some instances, by another FGC coordinator [11, p.22].

#### A4 Involvement of an interpreter at the Conference

In our base case scenario we exclude an interpreter, but an interpreter is included in analysis of an alternative scenario (Scenario C). Given the network-led decision-making process inherent in FGC services, it is essential that the FGC is conducted in the network’s preferred language wherever possible.

#### A5 Transport

We assume transportation is required for the FGC coordinator in order to visit the person and their network members in-person during the preparation stage and to attend the Conference. Our advising FGC practitioners emphasized the importance of in-person visits, as they are crucial for effectively promoting the idea of FGC. We assume that the LA/NHS covers travel costs for all professional participants, but not routinely for the central person and their network members. Exceptions are made for low-income families, where the social worker has discretion to provide a travel card or taxi pass.

#### A6 Food

Food and refreshments are assumed to be provided to all Conference attendees, at a cost to the LA/NHS; our preliminary work emphasised the importance of this, although we do present a scenario excluding food (Scenario D).

#### A7 Venue

The Conference is assumed to take place in a neutral venue chosen or agreed upon by the network (such as a community centre); alternatively, it may occur in the home of the person or a network member. Where the conference takes place at a venue other than a private residence (which is assumed to be the most common case), costs will be borne by the LA/NHS.

#### A8 Number of Network Members involved

We assume that three network members (e.g., family and friends) attend the Conference, based on the average number of participants reported by our advising FGC practitioners, reflecting practice in their organisations.

#### A9 Time involvement of service user and network members

We assume the central person spends two hours in one-to-one conversation with the FGC coordinator at the Preparation stage. We were advised that the FGC coordinator spends approximately one hour with each network member to share key information. We assume both the central person and network members attend the full Conference (the duration of which is four hours) and meetings occurring at the Review stage (two hours).

Resource use at each stage of the FGC process, and by each perspective, is reported in Table 1. Core elements (which would be expected in every typical good practice case) are presented with plain text formatting. Elements which may or may not be observed (such as inclusion of an advocate) are included in italics.

**Table 1:** Resource use by FGC stages and stakeholder perspectives.

| Types of Resource |  | Stage |  |  |
| --- | --- | --- | --- | --- |
|  |  | Preparation | Conference | Review |
| **LA/NHS perspective** |  |  |  |  |
| Staff<br>(hours worked) | FGC coordinator | 10 hours | 4 hours | 2 hours |
|  | Professional 1 | 0.5 hour | 4 hours | / |
|  | Professional 2 | 0.5 hour | 4 hours | / |
|  | *Advocate (optional)* | *2 hours* | *4 hours* | *2 hours* |
|  | *Interpreter (optional)* | *2 hours* | *4 hours* | *2 hours* |
| Non-staff | Travel | Coordinator visits the person and 3 x network members | Public transport costs for coordinator, referrer, and professionals | / |
|  | Food (and refreshments) | / | Catering for all participants during the Conference | / |
|  | Venue | / | Rental for 4 hours | / |
| **Central person's perspective** |  |  |  |  |
| Service user's time |  | 2 hours | 4 hours | 2 hours |
| Travel |  | / | Public transport costs | / |
| **Network members' perspective** |  |  |  |  |
| Network members' time (*3) |  | 1 hour per person | 4 hours per person | 2 hours per person |
| Travel |  | / | Public transport costs | / |

**Notes:** “/” denotes not applicable (no resource use). *Italic text* denotes resource use additional to that reflecting a typical case.

In total, we identified five scenarios that represent various possible and plausible FGC pathways in adult social care and mental health settings. The five scenarios are also summarised in Table 2.

- **Scenario A:** A full FGC pathway, incorporating all elements denoted in Table 1 with plain text formatting.
- **Scenario B:** A full FGC pathway, similar to Scenario A, but with the addition of an advocate at the Preparation, Conference and Review stages.
- **Scenario C:** A full FGC pathway, similar to Scenario A, but with the addition of an interpreter at the Preparation, Conference and Review stages.
- **Scenario D:** A full FGC pathway, similar to Scenario A, but excluding food provision at the Conference stage.
- **Scenario E:** A non-Conference (incomplete) FGC pathway, where the Conference does not take place. This may be because the central person and network members find that, as a result of the conversations initiated during the preparation phase, they can resolve their difficulties informally. Alternatively, a Conference may not proceed due to reasons such as the lack of a viable network or a significant change in circumstances.

**Table 2:**
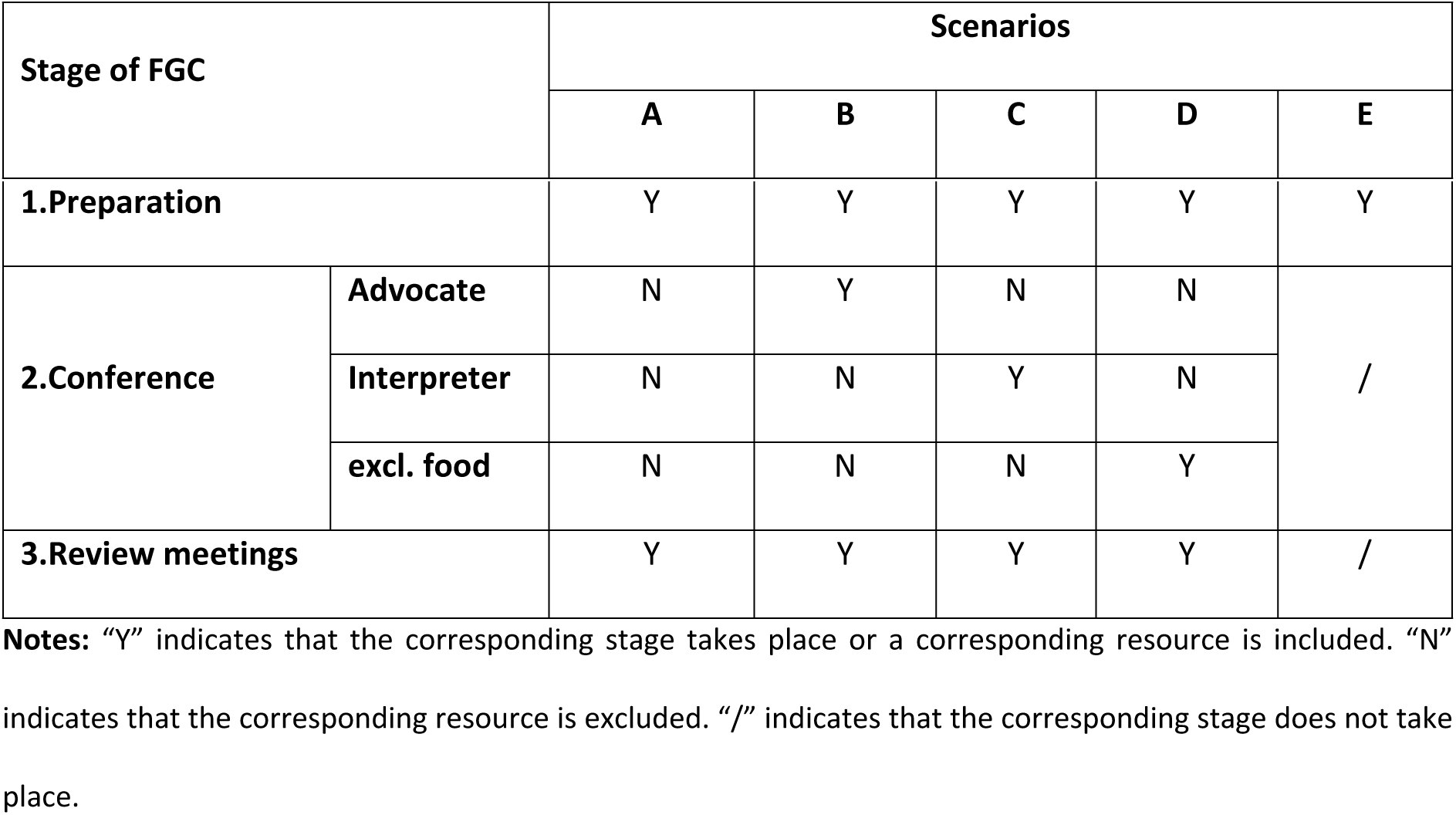
Five Scenarios of FGC pathways in adult social care/mental health settings (LA/NHS perspective)

| Stage of FGC | Scenarios |  |  |  |  |
| --- | --- | --- | --- | --- | --- |
|  | A | B | C | D | E |

|  |  |  |  |  |  |  |
| --- | --- | --- | --- | --- | --- | --- |
| **1.Preparation** |  | Y | Y | Y | Y | Y |
| **2.Conference** | **Advocate** | N | Y | N | N | / |
|  | **Interpreter** | N | N | Y | N |  |
|  | **excl. food** | N | N | N | Y |  |
| **3.Review meetings** |  | Y | Y | Y | Y | / |

**Notes:** “Y” indicates that the corresponding stage takes place or a corresponding resource is included. “N” indicates that the corresponding resource is excluded. “/” indicates that the corresponding stage does not take place.

**Table 3.**
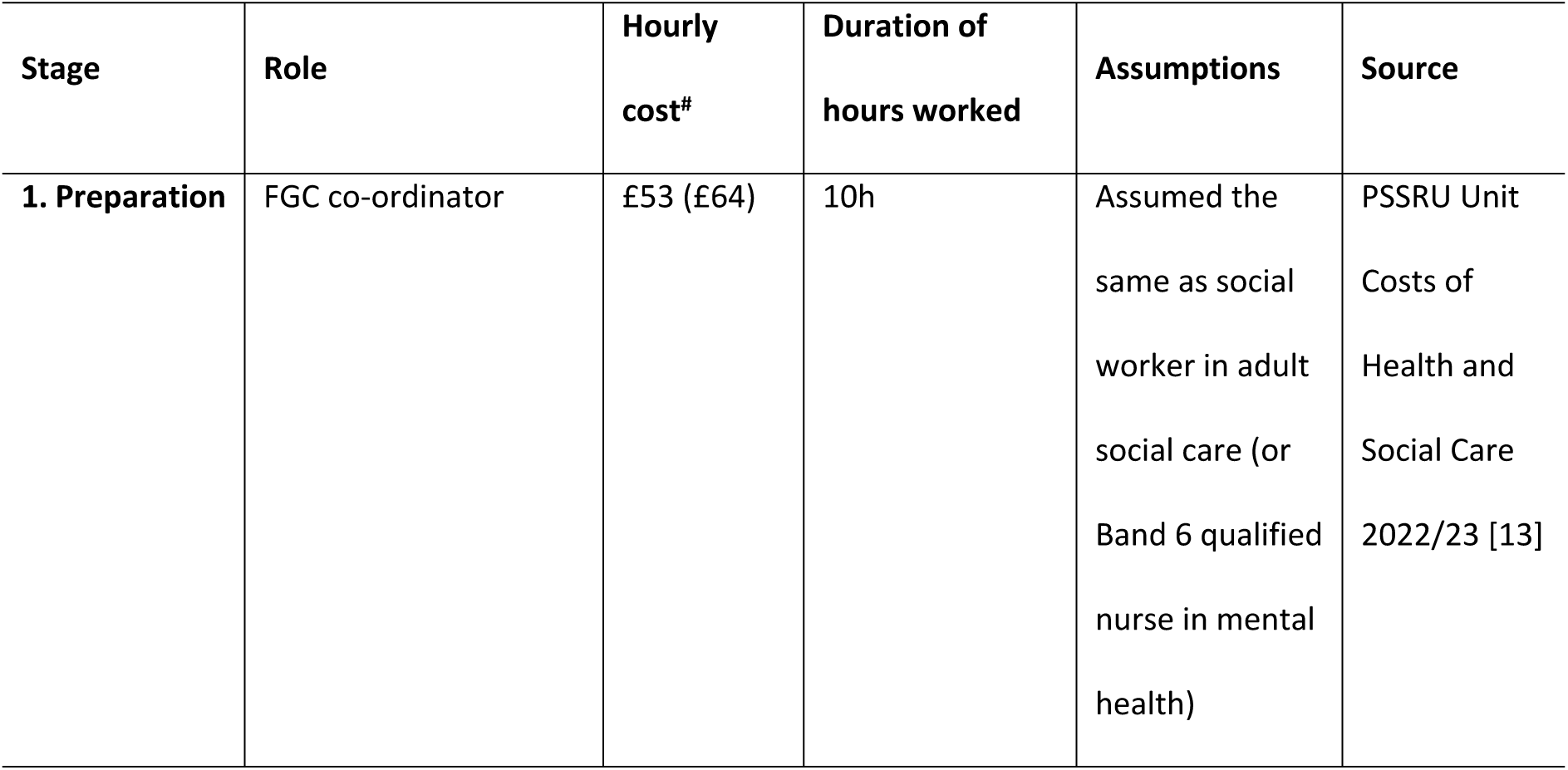
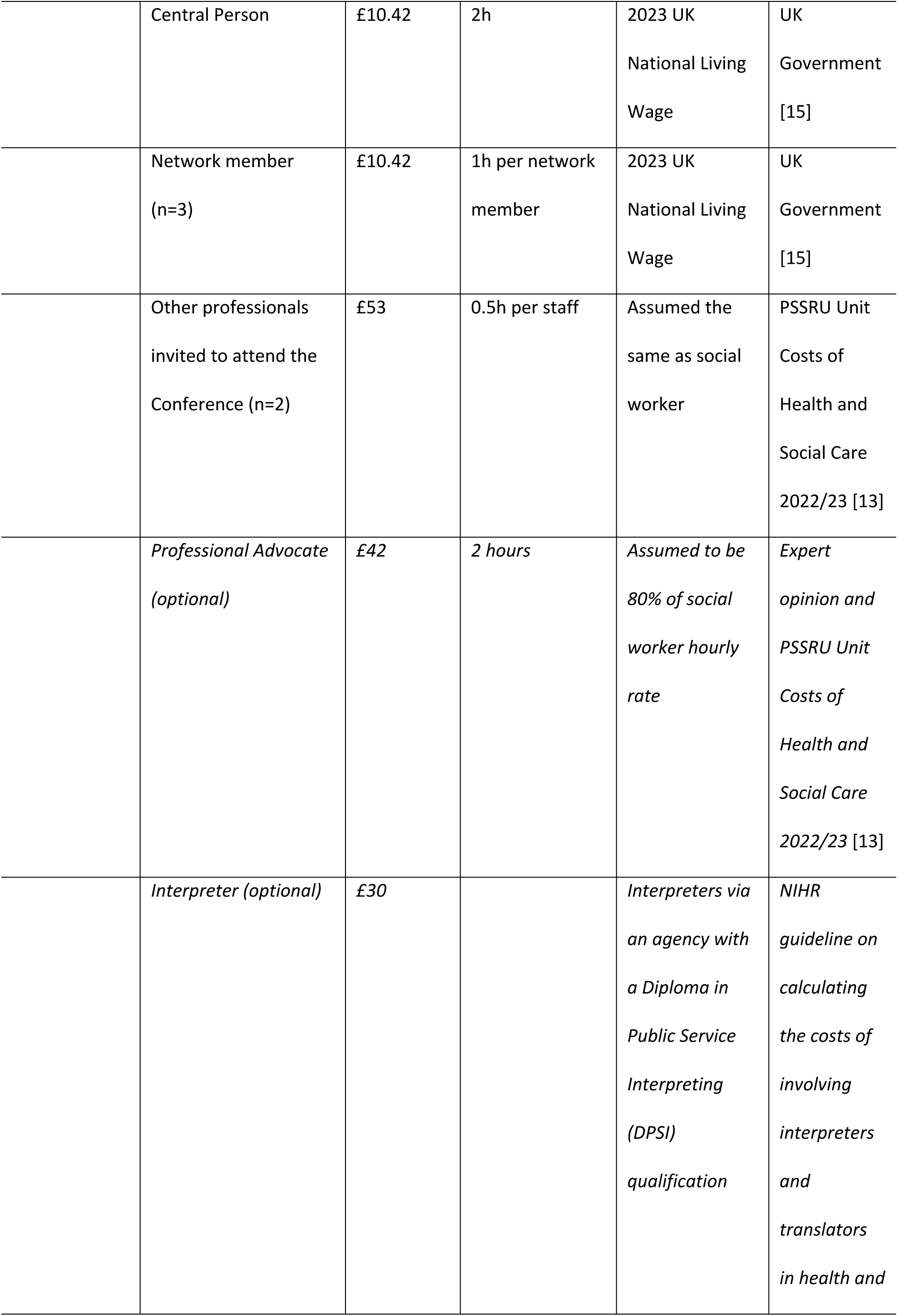
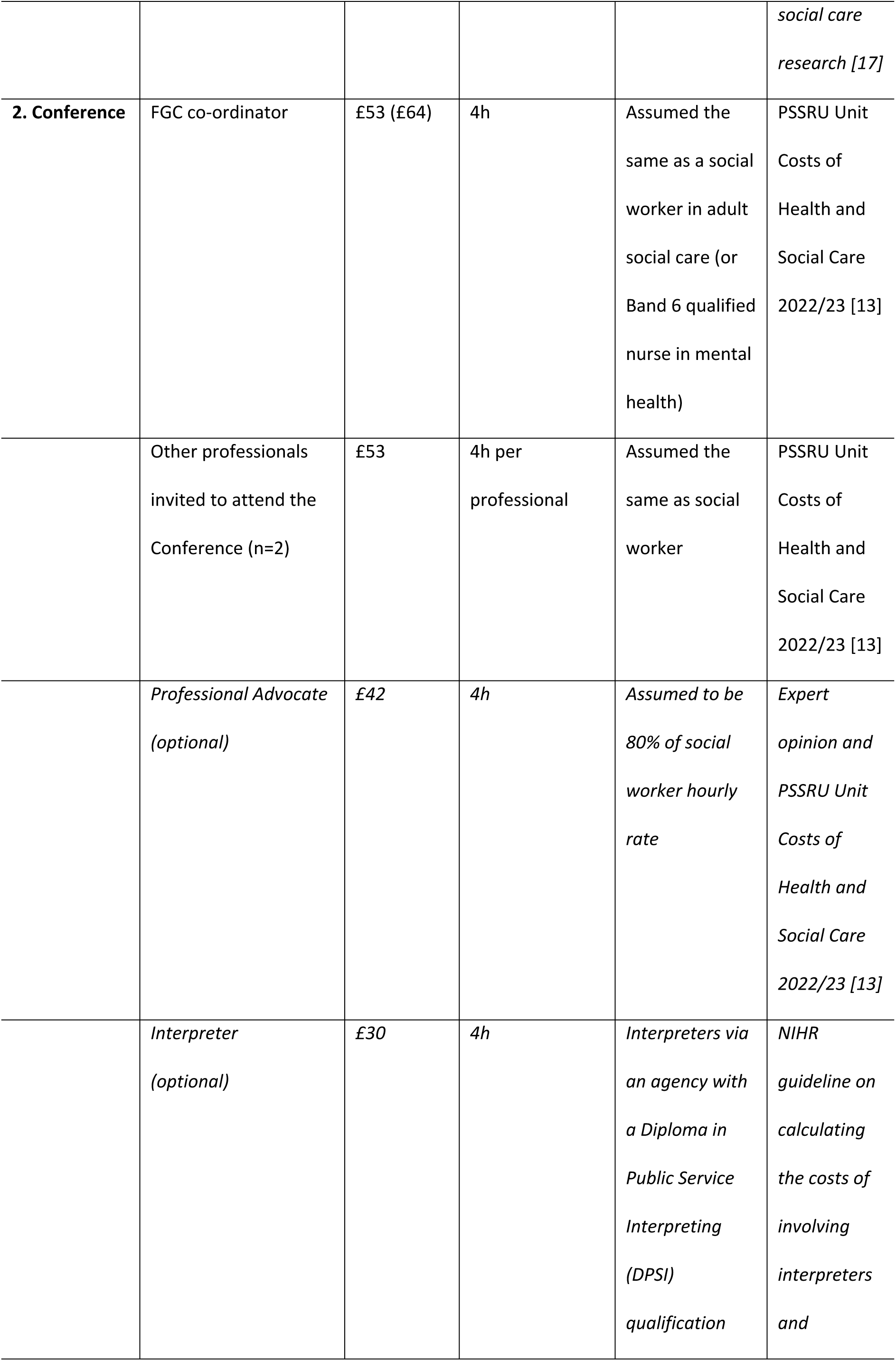
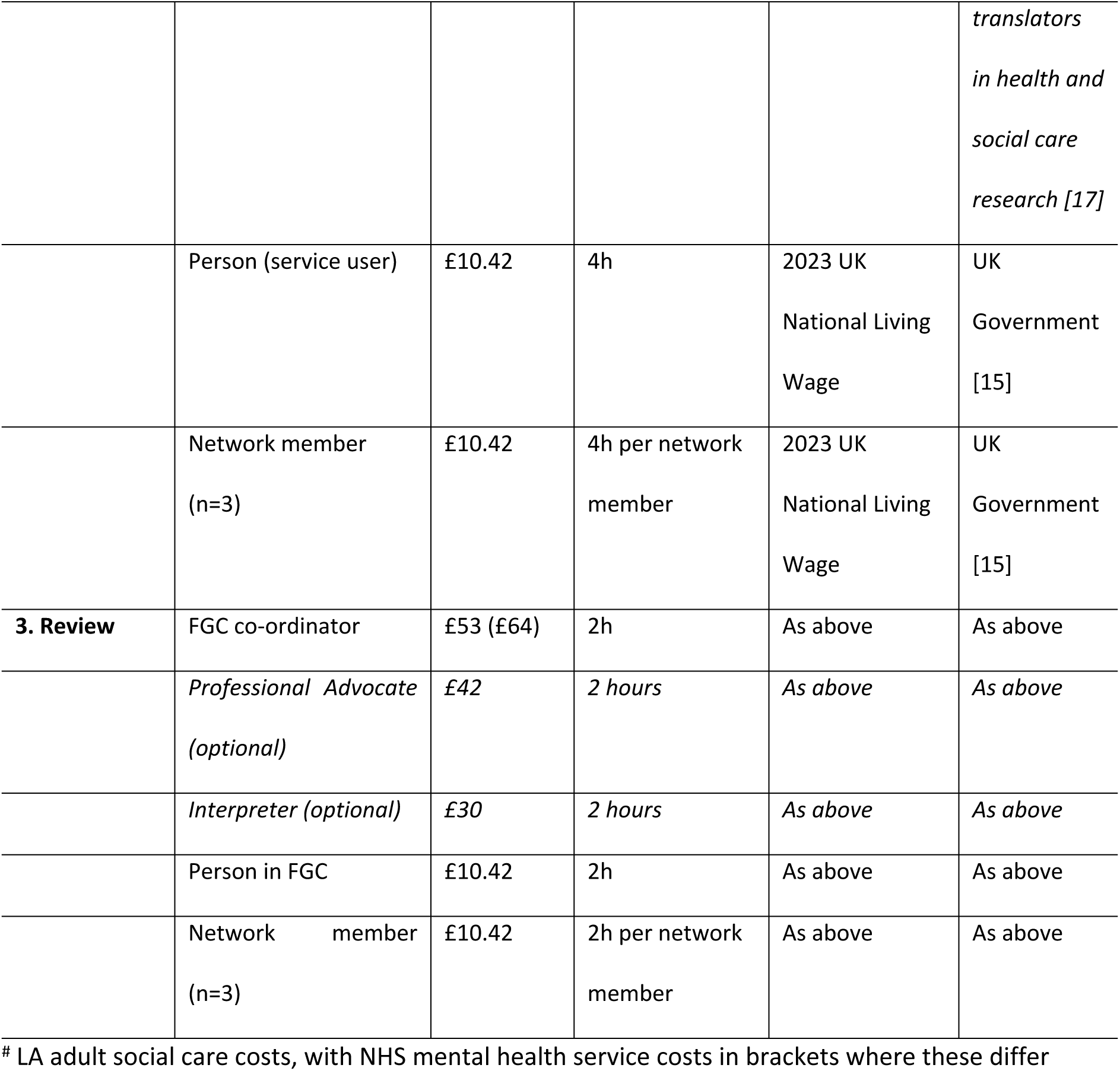
Hourly cost for all staff and participants’ time by stages of a full FGC.

**Table 4.**
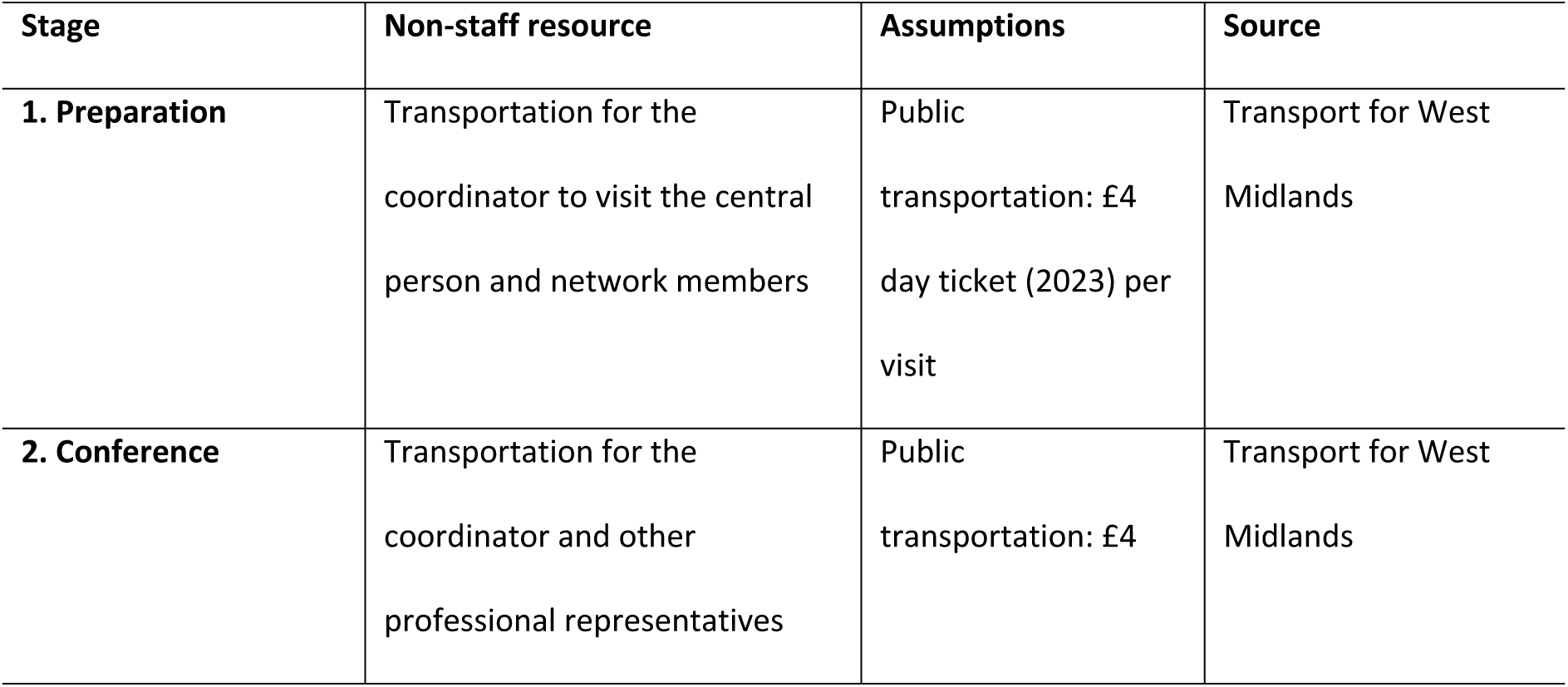
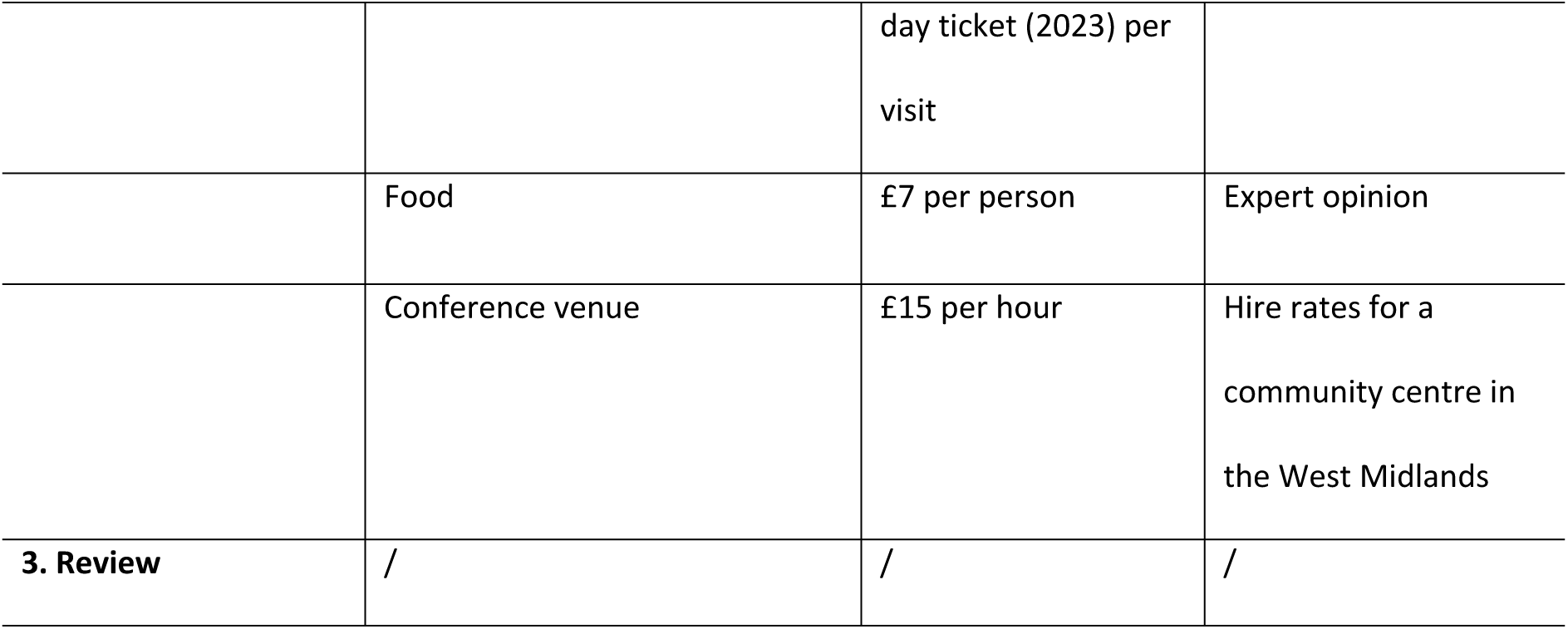
Non-staff Costs by stages of a full FGC (LA/NHS perspective)

There should be no systematic differences in resource use across scenarios A to D from a central person or network member perspective (i.e. resources they directly fund). Scenario E involves less time involvement for central persons, network members and professionals alike.

### 3.3. Monetary costs

Unit prices associated with all possible occurrences of resource use (documented in Table 1) are presented in Tables 3 (for staffing and personal time) and 4 (for non-staff/non-time costs), along with the source of that unit price.

A detailed cost breakdown associated with the three stages of our base case scenario (Scenario A) -calculated by multiplying quantities of resource use by unit costs-are reported in Table 5.

**Table 5.** Staff and non-staff costs of an FGC pathway in Scenario A (LA/NHS perspective)

| Stage of FGC | *Scenario A* |  |  |  |  |  |
| --- | --- | --- | --- | --- | --- | --- |
|  | *Adult Social Care* |  |  | *Mental health* |  |  |
|  | Staff costs<br>(£) | Non-staff costs (£) | Total (£) | Staff costs (£) | Non-staff costs (£) | Total (£) |
| **1.Preparation** | 583 | 16 | 599 | 693 | 16 | 709 |
| **2.Conference** | 636 | 121 | 757 | 680 | 121 | 801 |
| **3.Review** | 106 | 0 | 106 | 128 | 0 | 128 |
| **Total (£)** | 1325 | 137 | 1462 | 1501 | 137 | 1638 |

**Notes:** The differences between adult social care and mental health settings are due to the unit cost of the FGC coordinator differing across these two settings.

Next, we explore how costs vary across different scenarios (Table 6). The base case cost of a full FGC process in adult social care is £1,462 (see Panel A). The cost increases by 21% with the addition of an advocate and 17% with the addition of an interpreter. The increase is mainly driven by the added staff costs for these roles. Excluding food reduces the cost by 3%. As a result, the total cost ranges from £1,413 to £1,773. If the FGC does not proceed to the Conference stage, the cost drops to £599, which is 41% of the baseline. In the mental health setting (see Panel B), a full FGC pathway is more expensive, ranging from £1,589 to £1,985. If the Conference stage is not reached, the cost is reduced to £709.

**Table 6.**
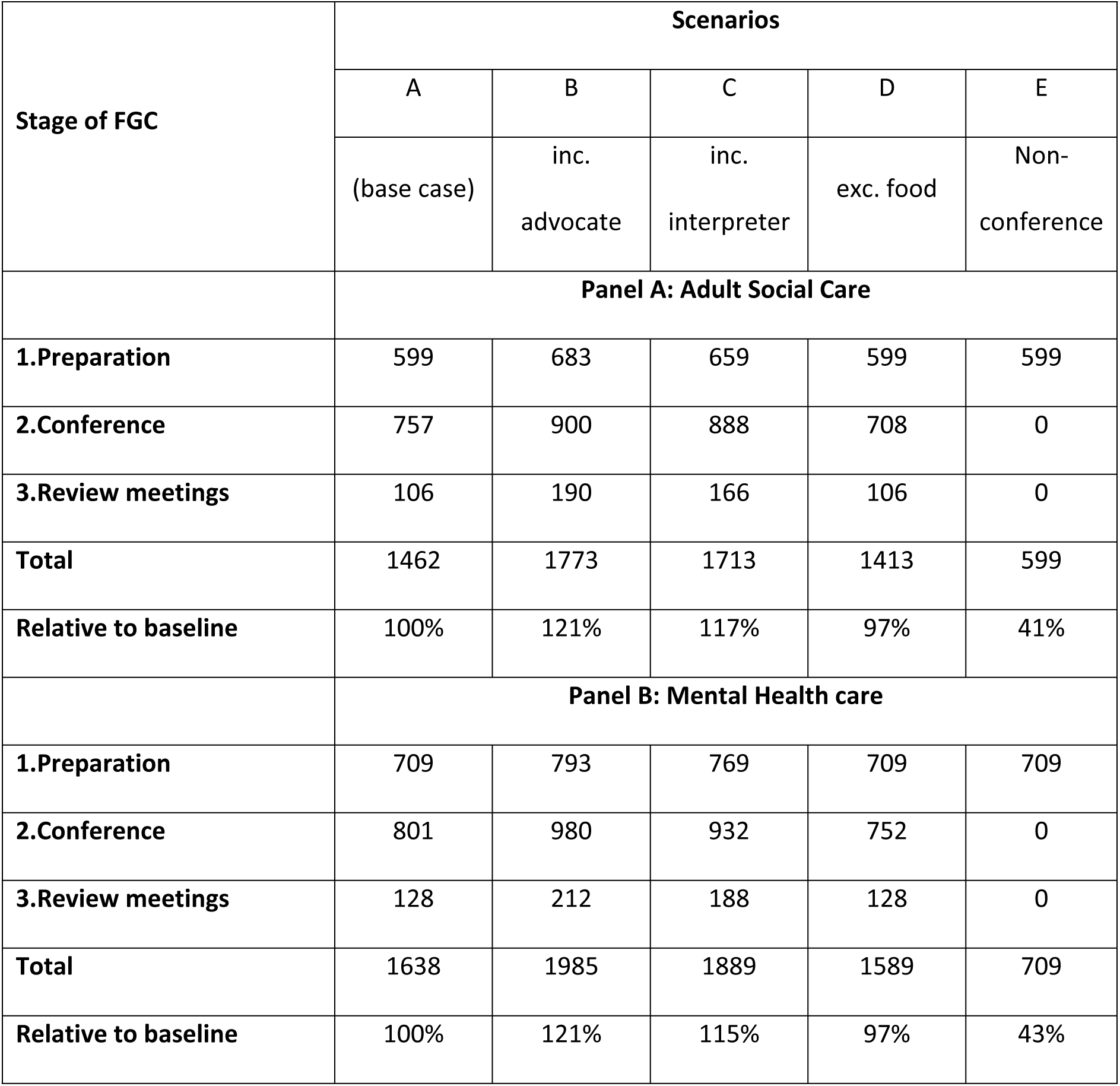
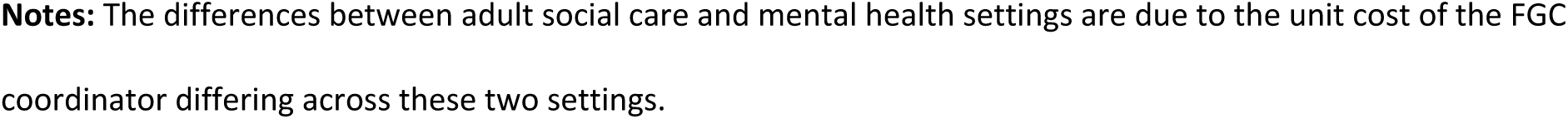
Costs of FGC pathways in five scenarios (LA/NHS perspective)

| Stage of FGC | Scenarios |  |  |  |  |
| --- | --- | --- | --- | --- | --- |
|  | A | B | C | D | E |
|  | (base case) | inc.<br>advocate | inc.<br>interpreter | exc. food | Non-conference |
|  | **Panel A: Adult Social Care** |  |  |  |  |
| **1.Preparation** | 599 | 683 | 659 | 599 | 599 |
| **2.Conference** | 757 | 900 | 888 | 708 | 0 |
| **3.Review meetings** | 106 | 190 | 166 | 106 | 0 |
| **Total** | 1462 | 1773 | 1713 | 1413 | 599 |
| **Relative to baseline** | 100% | 121% | 117% | 97% | 41% |
|  | **Panel B: Mental Health care** |  |  |  |  |
| **1.Preparation** | 709 | 793 | 769 | 709 | 709 |
| **2.Conference** | 801 | 980 | 932 | 752 | 0 |
| **3.Review meetings** | 128 | 212 | 188 | 128 | 0 |
| **Total** | 1638 | 1985 | 1889 | 1589 | 709 |
| **Relative to baseline** | 100% | 121% | 115% | 97% | 43% |

**Notes:** The differences between adult social care and mental health settings are due to the unit cost of the FGC coordinator differing across these two settings.

**Table 7.**
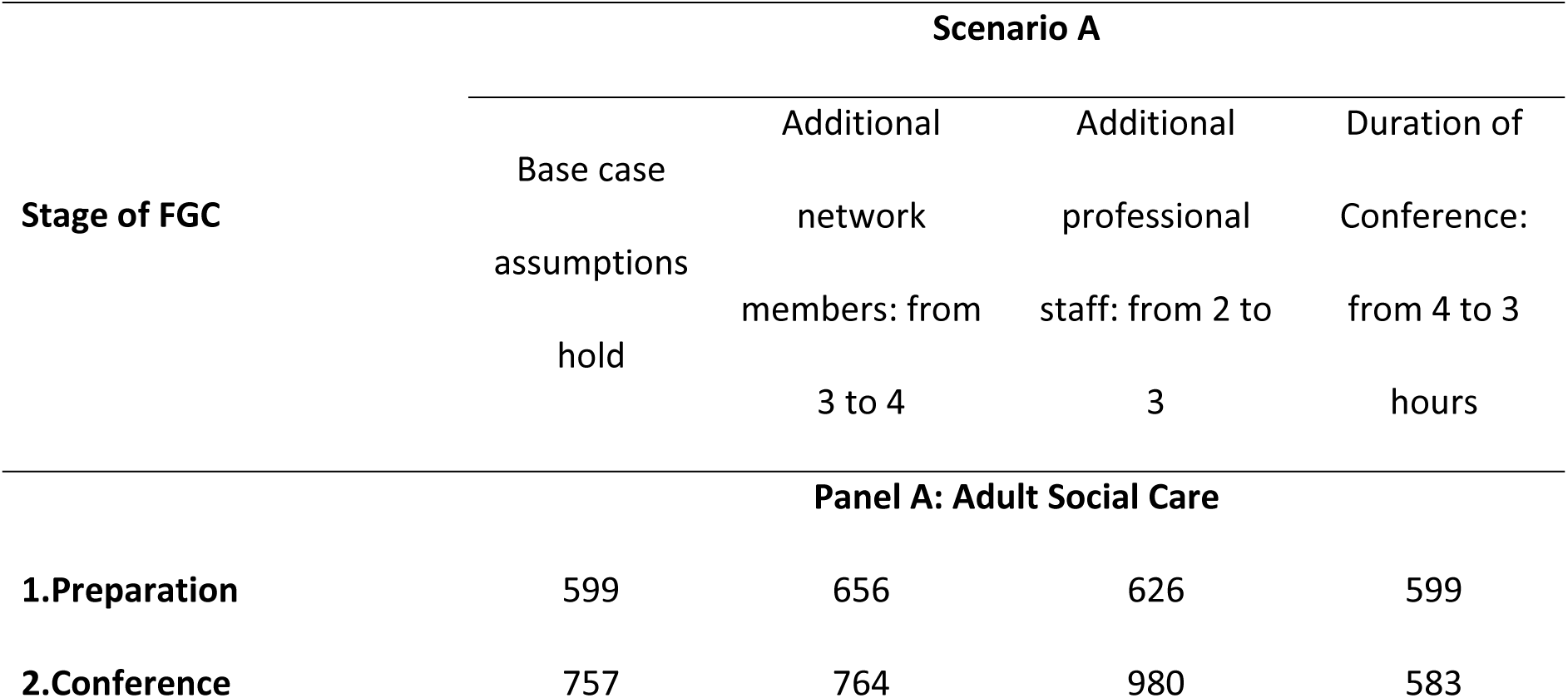
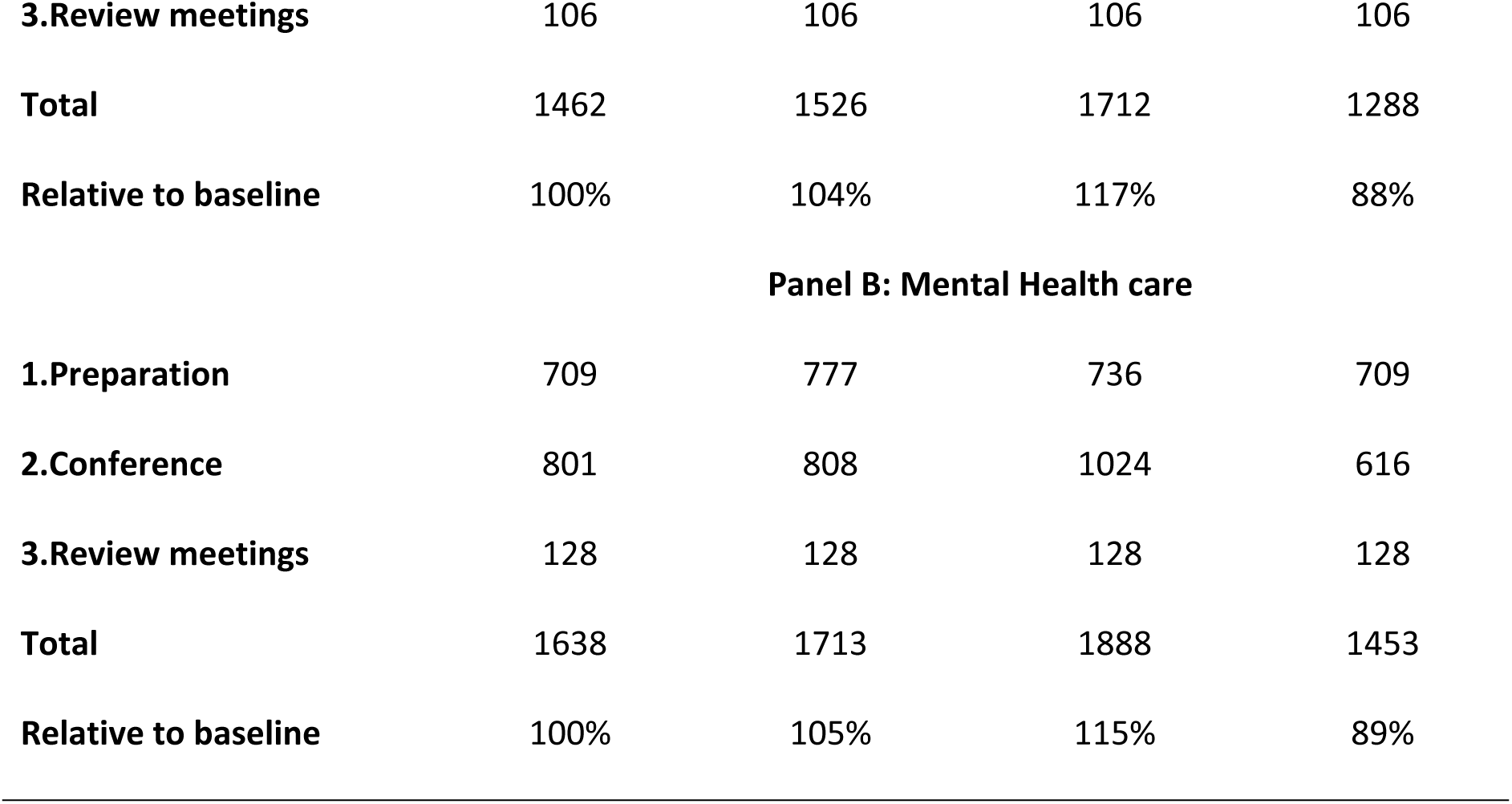
Sensitivity results (LA/NHS perspective)

| Stage of FGC | Scenario A |  |  |  |
| --- | --- | --- | --- | --- |
|  | Base case | Additional | Additional | Duration of |
|  | assumptions | network | professional | Conference: |
|  | hold | members: from | staff: from 2 to | from 4 to 3 |
|  |  | 3 to 4 | 3 | hours |
| Panel A: Adult Social Care |  |  |  |  |
| 1.Preparation | 599 | 656 | 626 | 599 |
| 2.Conference | 757 | 764 | 980 | 583 |
| **3.Review meetings** | 106 | 106 | 106 | 106 |
| **Total** | 1462 | 1526 | 1712 | 1288 |
| **Relative to baseline** | 100% | 104% | 117% | 88% |
| **Panel B: Mental Health care** |  |  |  |  |
| **1.Preparation** | 709 | 777 | 736 | 709 |
| **2.Conference** | 801 | 808 | 1024 | 616 |
| **3.Review meetings** | 128 | 128 | 128 | 128 |
| **Total** | 1638 | 1713 | 1888 | 1453 |
| **Relative to baseline** | 100% | 105% | 115% | 89% |

**Table 8.** Costs of FGC pathways in two scenarios for central person and each network member.

| Stage of FGC | Central person perspective |  | Per network member perspective |  |
| --- | --- | --- | --- | --- |
|  | Scenario A | Scenario E | Scenario A | Scenario E |
| **1.Preparation** | 20.84 | 20.84 | 10.42 | 10.42 |
| **2.Conference** | 45.68 | NA | 45.68 | NA |
| **3.Review** | 20.84 | NA | 20.84 | NA |
| **Total (£)** | 87.36 | 20.84 | 76.94 | 10.42 |

**Notes:** NA = Not Applicable.

Adjusting to March 2025 prices, using the Bank of England inflation calculator [16], gives an adult social care base case (Scenario A) cost of £1,505, and an NHS Mental Health base case cost of £1,686. Scenario B costs increase to £1,825 and £2,043 respectively. Scenario C costs at March 2025 prices are £1,763 and £1,945 respectively. Scenario D £1,455 and £1,636. Scenario E £617 and £730.

### 3.4 Sensitivity results

In this section we adjust the assumptions made earlier to explore how changes in the number of network members, professional staff attending, and the duration of the Conference affect the total base case cost (Scenario A in Table 6). The first column presents the base case costs from Table 6 for reference. The results for adult social care and mental health care settings are presented separately in Panel A and Panel B, respectively.

From a LA/NHS perspective, adding an extra network member increases the total cost from £1,462 to £1,1526 an increase of £64. This relatively minor increase is due to additional time spent by the FGC coordinator at the preparation stage (1 additional hour), additional travel costs incurred by the FGC Coordinator during the preparation stage and additional food costs at the conference. Adding an additional professional member, however, increases the total cost from £1,462 to £1,712, an increase of £250. This 17% increase from base case cost is primarily due to the additional staff expense, along with the added costs for food and transportation for the extra attendee. Reducing the duration of the Conference can lead to substantial cost saving. From a LA perspective, shortening the Conference to 3 hours decreases the total cost from £1,462 to £1,288. This reduction of £174, or 12% of the baseline cost is primarily due to the reduced hours worked of the coordinator, and practitioners invited or required. These effects are similar in a mental health care setting as presented in Panel B.

Finally, we acknowledge the voluntary nature of FGC services, recognizing that some families might choose not to proceed to the Conference stage. We have made assumptions about the probability that an FGC referral will advance to the Conference stage and present our estimates accordingly. Scenario E is presented as the non-Conference pathway (see Appendix Table A1). If 65% of FGC referrals result in a full FGC pathway (Scenario A) and 35% result in a non-Conference pathway (Scenario E), the expected cost per case *accepted* into the FGC service for adult social care would be approximately £1,160, and £1,313 in NHS mental health services.

### 3.5 Cost from other stakeholder perspectives

We present the costs for the central person and each network member across the two potential FGC pathways that are distinct from their perspective (see Table). During the preparation stage, we assumed the central person spent more time with the FGC coordinator, so the associated cost from the central person perspective is higher.

### 3.6 Inclusion of referral stage in FGC costs

The referral stage is a prerequisite step for FGC [1], which adds cost associated with both the referrer and the FGC manager (the FGC coordinator and FGC manager are distinct and separate roles). This referral is typically made by professionals such as a social worker in adult social care or a care coordinator in mental health, who identifies a situation where FGC could be beneficial. The case is then reviewed by the FGC manager or team lead to determine if the referral criteria are met. If so, an FGC coordinator is appointed to facilitate the process.

Appendix Table A2 presents the costs including referral. Cost differences between the two settings are due to the assumed role of the referrer along with the unit cost of the FGC manager during the Referral. In adult social care, the total cost of a full FGC pathway, including referral, ranges from £1,569 to £1,879 (higher than the range excluding the referral stage: £1,462 to £1,773). Similarly, in mental health care setting, the total cost increases from £1,717 to £1,766 when the referral stage is included. Costs in the mental health care setting may rise further if the referrer is assumed to be a psychiatrist (a more senior clinical role). Feedback from our advising FGC practitioner in mental health care suggests that whilst referrals have historically come from care coordinators, there has been a growing number of referrals directly from psychiatrists. To account for this, costs were also calculated assuming the referrer to be a hospital-based psychiatric consultant, with an hourly cost of £143 (2022/23 rate with qualifications). The corresponding total costs, reported in Appendix Table A3, show that an FGC pathway including referral is £1,845 for scenario A. If the Conference stage is not reached, the cost is reduced to £916.

## 4 Discussion

In this study, we aimed to explore the costs associated with Family and Group Conferencing (FGC) pathways in adult social care and mental health settings from multiple stakeholder perspectives using a mixed methods approach, drawing on data from the literature, programme theory developed in advance of our costing work, and expert opinion. Our findings indicate that the costs of conducting a full FGC (in 2022/23 prices) range from £1,413 to £1,985 depending on factors such as the provision of food and a venue, the involvement of an advocate or interpreter, and the setting (adult social care or mental health), but excluding the Referral stage. Adjusted to March 2025 prices, the range is £1,455 to £2,043.

We conducted sensitivity analyses to examine the impact of having additional network members, professional staff and varying the duration of the Conference on the overall cost. The inclusion of an additional network member results in only a marginal increase (4-5%) in the overall cost, but the involvement of an additional professional can increase cost by 15-17%. Reducing the Conference duration (from four hours to three hours) could decrease the total cost by 11-12%.

Recognizing that some families may opt not to proceed to the Conference stage, we also estimated the expected cost per case accepted into the FGC service, adjusting for the probability of proceeding to the Conference and a full FGC. The expected cost per case accepted into an FGC service for adult social care would be approximately £1,160, and £1,313 in NHS mental health services in 20222/23 prices (£1,194 and £1,352, respectively, when adjusted to March 2025 prices). We also estimated the cost of engaging in FGC for the central person and each network member. Separate results were presented for FGC pathways that include referral stage, where additional costs arise due to referrer’s and FGC manager’s time.

This is the first bottom-up costing of FGC in the contexts of adult social care and NHS Mental Health Services. The strength of the bottom-up costing is its foundation in the programme theory, developed in earlier phases of the project, the use of expert practitioner opinion to ensure that each assumption reflects practice in reality, use of credible cost sources (such as PSSRU Unit Costs of Health & Social Care), and the scenario-based analysis, including sensitivity analysis. However, the study has some limitations. We utilized data from three experts, representing three established FGC services, but there may be variation in FGC implementation across different services that were not fully captured. Some FGC services are shifting from in-person to online formats, a trend that has accelerated during the pandemic [18]. This transition could reduce overall costs by saving on travel, food and venue hire. However, this cost reduction might come at the expense of the trust built between the network and professionals, which may be harder to establish in a virtual setting. Furthermore, we have assumed the professional staff engage fully at the Conference for simplicity. However, in practice, their level of involvement may vary.

These cost estimates highlight the financial investment required by LA/NHS to provide an FGC in adult social care and mental health settings. The inclusion of additional support personnel, such as advocates/interpreters and professionals, as well as the provision of food and venue, increases the overall cost. Furthermore, extended durations at the Conference stage can escalate costs, although, in principle, families are allowed unlimited time to formulate and agree upon a plan [1]. Whilst FGC services are a valuable tool that local authorities can use to meet their requirements under the Care Act, the investment in FGC must be justified in terms of improved outcomes, and full evidence on economic effectiveness.

Although current evaluations of FGC studies primarily draw from the child welfare context, our cost estimates align with broader literature on FGC costs across various settings. For instance, a rapid evidence review conducted in Camden noted that a standard FGC in early help settings for children typically costs the LA between £1,200 and £1,500, excluding the referral process [19]. Additionally, a 2015-2016 evaluation in Leeds reported the cost of providing FGC services as part of a broader initiative to implement restorative practices in children’s services at £2,418 per family [9]. These variations in cost estimates can be attributed to different methodological approaches, study sites, the specific needs of the central person and the professionals involved.

Policymakers and future research should consider the financial cost of FGC alongside the potential for future cost-saving. However, policy-makers and service providers should not focus solely cost-saving measures without accounting for outcomes. The cost estimates presented in this paper are a preliminary step towards a comprehensive economic evaluation of FGC services in adult social care and mental health, which will evolve as more outcome data becomes available.

A critical challenge in conducting full economic evaluation is selecting an appropriate counterfactual scenario – typically, “services-as-usual”. If the counterfactual is not accurately chosen, analysis may misrepresent the costs and benefits of FGC services, potentially leading to incorrect conclusions about its value.

## Data Availability

There are no transcripts, field notes or datasets. All assumptions and sources are specified or referenced within the paper. Data for the papers by Mahesh, S (2025) and Tew, J (2025) which set out the programme theory are both open access and data underpinning those papers is publicly accessible.

## Acknowledgements

We would like to thank staff across our three case study sites for sharing their knowledge and experience and hence informing key assumptions and parameters underpinning our costings.

